# Angiographic Report Generation for the 3^rd^ APTOS’s Competition: Dataset and Baseline Methods

**DOI:** 10.1101/2023.11.26.23299021

**Authors:** Weiyi Zhang, Peranut Chotcomwongse, Xiaolan Chen, Florence H.T. Chung, Fan Song, Xueli Zhang, Mingguang He, Danli Shi, Paisan Ruamviboonsuk

## Abstract

Fundus angiography, including fundus fluorescein angiography (FFA) and indocyanine green angiography (ICGA), are essential examination tools for visualizing lesions and changes in retinal and choroidal vasculature. However, the interpretation of angiography images is labor-intensive and time-consuming. In response to this, we are organizing the third APTOS competition for automated and interpretable angiographic report generation. For this purpose, we have released the first angiographic dataset, which includes over 50,000 images labeled by retinal specialists. This dataset covers 24 conditions and provides detailed descriptions of the type, location, shape, size and pattern of abnormal fluorescence to enhance interpretability and accessibility. Additionally, we have implemented two baseline methods that achieve an overall score of 7.966 and 7.947 using the classification method and language generation method in the test set, respectively. We anticipate that this initiative will expedite the application of artificial intelligence in automatic report generation, thereby reducing the workload of clinicians and benefiting patients on a broader scale.

## Introduction

Fundus fluorescein angiography (FFA) and indocyanine green angiography (ICGA) are prevalent and essential diagnostic imaging tests in clinical ophthalmology. They are particularly useful in identifying abnormal changes in the retinal and choroidal vasculature, thereby facilitating the diagnosis and monitoring of eye diseases such as retinal vascular occlusion, diabetic retinopathy, and central serous chorioretinopathy through the dynamic changes of injected dyes.[1-3] Given that a patient’s angiographic images can range from tens to hundreds, their interpretation can be labor-intensive and prone to errors.

The advent of artificial intelligence (AI) has sparked a trend towards the automatic interpretation of medical reports.[4-6] The automatic generation of ophthalmic angiography reports could significantly alleviate the workload of ophthalmologists.[7] Furthermore, these generated reports could potentially reduce the oversight of abnormalities and enhance the accuracy of retinal diagnoses.[8] Researchers have proposed cutting-edge methods that combine computer vision, natural language processing, and clinical knowledge to generate reliable medical reports.[9] Li et al. introduced a Cross-modal clinical Graph Transformer for generating FFA reports.[8] Chen et al. combined a vision transformer with a large language model for FFA report generation and question answering.[10] However, while medical report generation requires substantial and interpretable real-world data for training[11], there is a lack of publicly available FFA and ICGA datasets for report generation.[12]

To address these challenges, we have curated a large angiographic dataset that includes both FFA and ICGA images with interpretable labels and proposed baseline methods for angiographic report generation. The APTOS big data competition was organized to promote the development of medical report generation algorithms in the field of ophthalmic angiography. We anticipate that our approach will significantly expedite the application of AI in ophthalmology.

## Methods

### Dataset

We retrospectively collected 58,520 angiographic images from 1,711 patients (3,405 eyes) from the Department of Ophthalmology, Rajavithi Hospital, Bangkok, Thailand. One ophthalmologist (P.C.) with more than five years of clinical experience reviewed the images for each patient and labeled the images in regards to the impression of disease, and abnormal fluorescence pattern: the fluorescence type (hyper or hypo fluorescence), the location of fluorescence (in depth direction [retinal/subretinal], in x-direction [at the macula, disc or other], the size of abnormal fluorescence, the special pattern of fluorescence (polyps, ink dot) and vascular abnormality due to diabetic retinopathy. Blurry mages that could not reach a diagnosis will be excluded. The time of the angiographic images was recognized by Optical Character Recognition (OCR)[13], and phases were categorized based on time. Where time < 25 seconds was categorized into arterial venous phase, time >=25 seconds and <5 minutes was categorized into venous phase, and time >=5 minutes was categorized into late phase. The characteristics of the dataset are illustrated in **Table 1**.

**Table 1.** Dataset characteristics.

|  | Total (n = 3179) |
| --- | --- |
| eye, n (%) |  |
| L | 1590 (50) |
| R | 1589 (50) |
| Hyperfluorescence Type, n (%) |  |
| leakage | 1255 (39.5) |
| no | 775 (24.4) |
| pooling | 312 (9.8) |
| staining | 632 (19.9) |
| window defect | 205 (6.4) |
| Hyperfluorescence Area(Disc Area), n (%) |  |
| 4 | 1946 (61.2) |
| 5 | 458 (14.4) |
| no | 775 (24.4) |
| Hyperfluorescence Fovea, n (%) |  |
| no | 1627 (51.2) |
| yes | 1552 (48.8) |
| Hypofluorescence Type, n (%) |  |
| blockage | 772 (24.3) |
| capillary non-perfusion | 120 (3.8) |
| no | 2287 (71.9) |
| Hypofluorescence Area(Disc Area), n (%) |  |
| 4 | 656 (20.6) |
| 5 | 239 (7.5) |
| no | 2284 (71.8) |
| Hypofluorescence Fovea, n (%) |  |
| no | 2543 (80) |
| yes | 636 (20) |
| CNV, n (%) |  |
| no | 2086 (65.6) |
| yes | 1093 (34.4) |
| Angiographic Images (n = 55361) |  |
| Examination mode, n (%) |  |
| FA&&ICGA | 45273 (81.8) |
| FA | 5702 (10.3) |
| ICGA | 4386 (7.9) |
| Phase, n (%) |  |
| Venous | 28987 (52.4) |
| Late | 24579 (44.4) |
| Arterial venous | 1786 (3.2) |
| Image shape, n (%) |  |
| (868, 1536) | 50157 (90.6) |
| (1636, 1536) | 3444 (6.2) |
| (868, 768) | 1465 (2.6) |
| (612, 1024) | 285 (0.5) |
| (1124, 1024) | 10 (0.0) |

B
| Impression | Count (Freq) |
| --- | --- |
| macular neovascularization | 1077 (32.5%) |
| unremarkable changes | 720 (21.8%) |
| dry age-related macular degeneration | 339 (10.2%) |
| central serous chorioretinopathy | 299 (9.0%) |
| uveitis | 153 (4.6%) |
| chorioretinal scar | 151 (4.6%) |
| diabetic retinopathy | 106 (3.2%) |
| retinal pigment epithelial detachment | 93 (2.8%) |
| polypoidal choroidal vasculopathy | 81 (2.4%) |
| pachychoroid pigment epitheliopathy | 70 (2.1%) |
| chorioretinal atrophy | 34 (1.0%) |
| myopia | 26 (0.8%) |
| proliferative diabetic retinopathy | 26 (0.8%) |
| cystoid macular edema | 23 (0.7%) |
| choroidal mass | 23 (0.7%) |
| other | 18 (0.5%) |
| epiretinal membrane | 16 (0.5%) |
| retinal vein occlusion | 15 (0.5%) |
| retinal arterial macroaneurysm | 9 (0.3%) |
| branch retinal vein occlusion | 8 (0.2%) |
| central retinal vein occlusion | 7 (0.2%) |
| retinal dystrophy | 6 (0.2%) |
| diabetic macular edema | 5 (0.2%) |
| central retinal artery occlusion | 4 (0.1%) |

The angiographic images were divided into train, validation, and test set at subject-level by 6:2:2, there was no patient overlap among different splits. **Figure 1** shows the workflow of the study. **Figure 2** shows the examples of images in different shapes.

**Figure 1.**
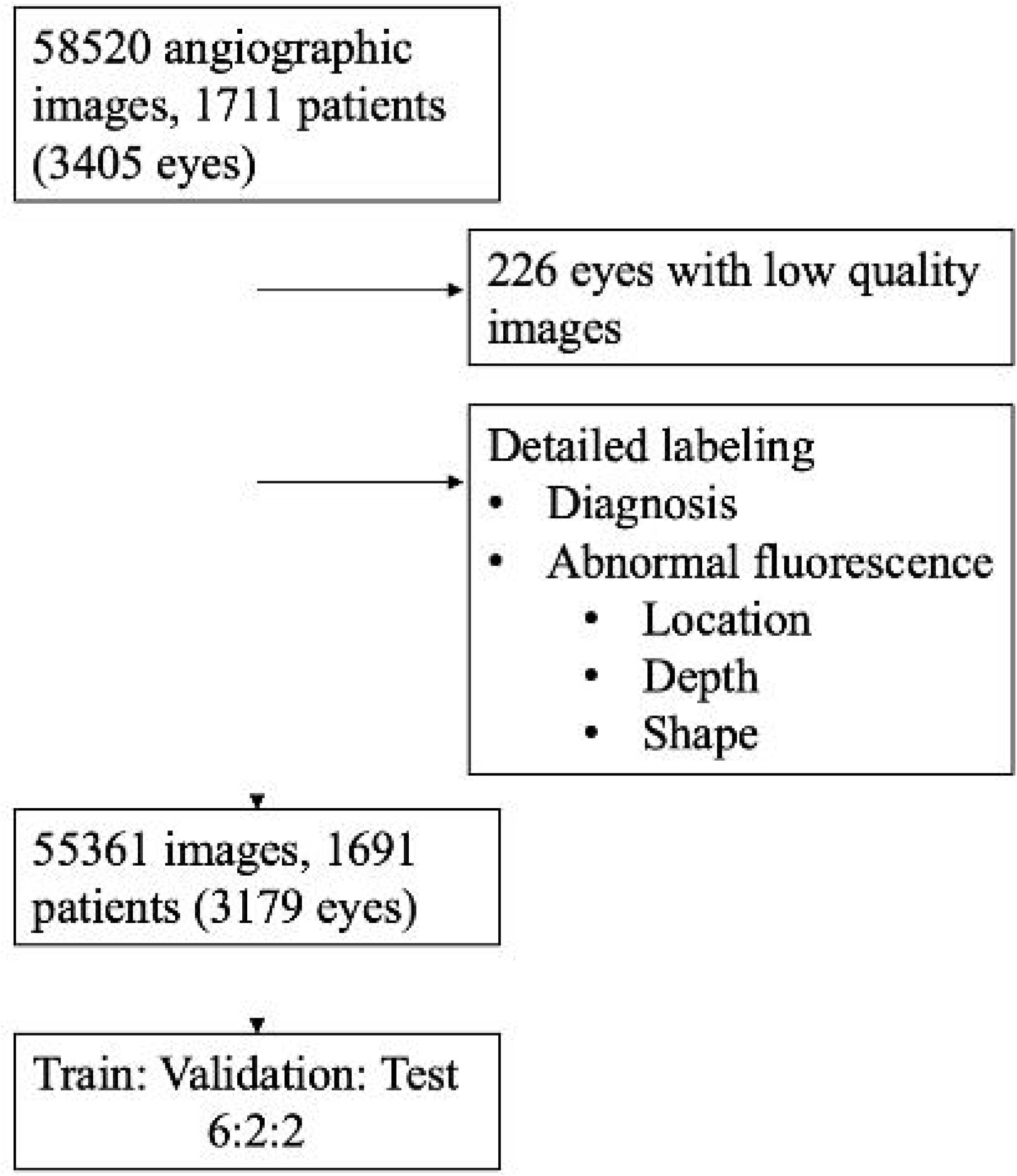
Workflow of the study.

**Figure 2.**
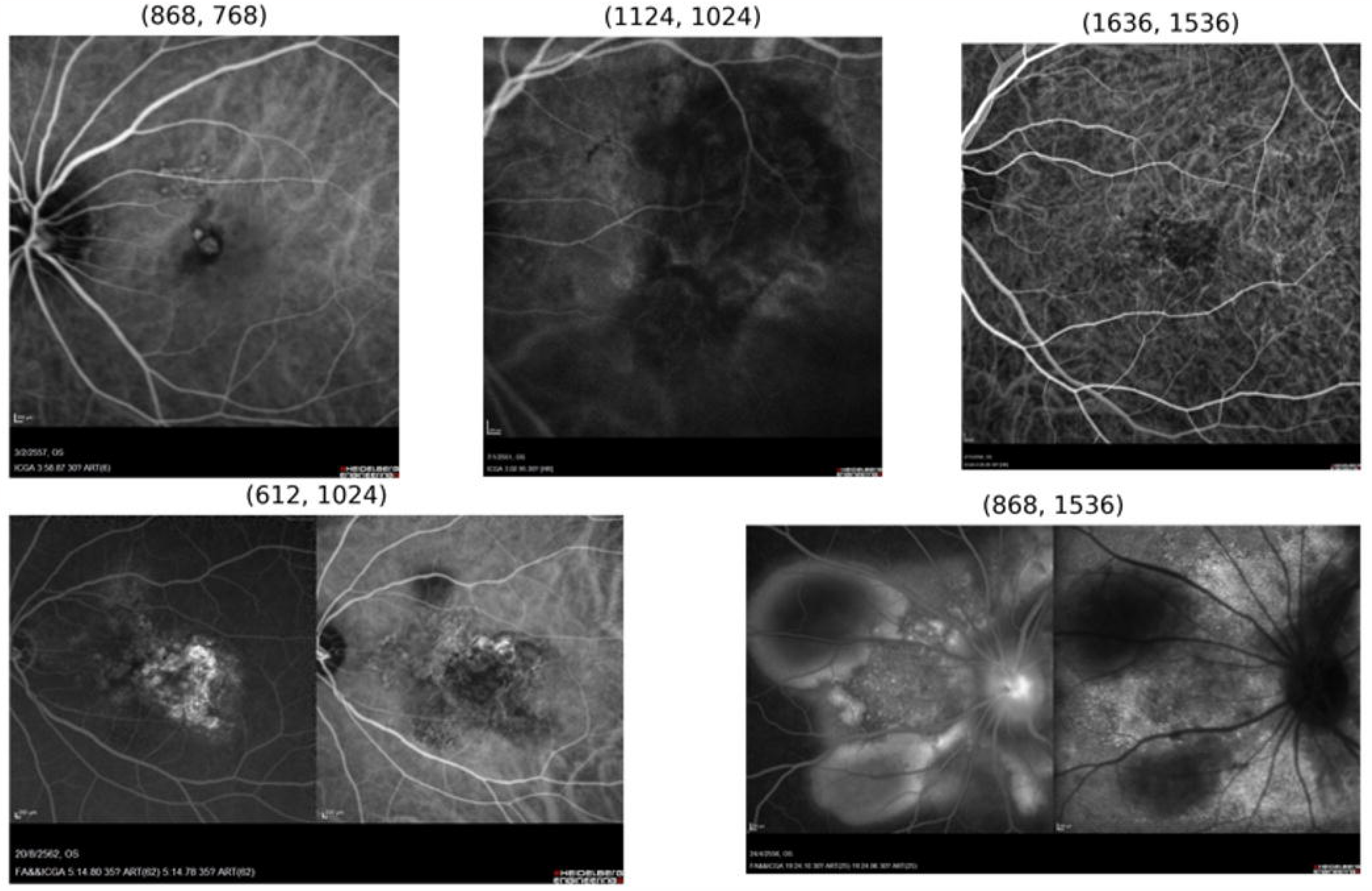
Examples of angiographic images in different sizes.

The images were de-identified and the retrospective study was approved by the institutional review board of Rajavithi Hospital.

### Baseline solutions

We employed both the classification and language-generation methods to investigate and compare their performances in our report-generation task comprehensively. The demonstration of different methods is shown in **Figure 3**.

**Figure 3.**
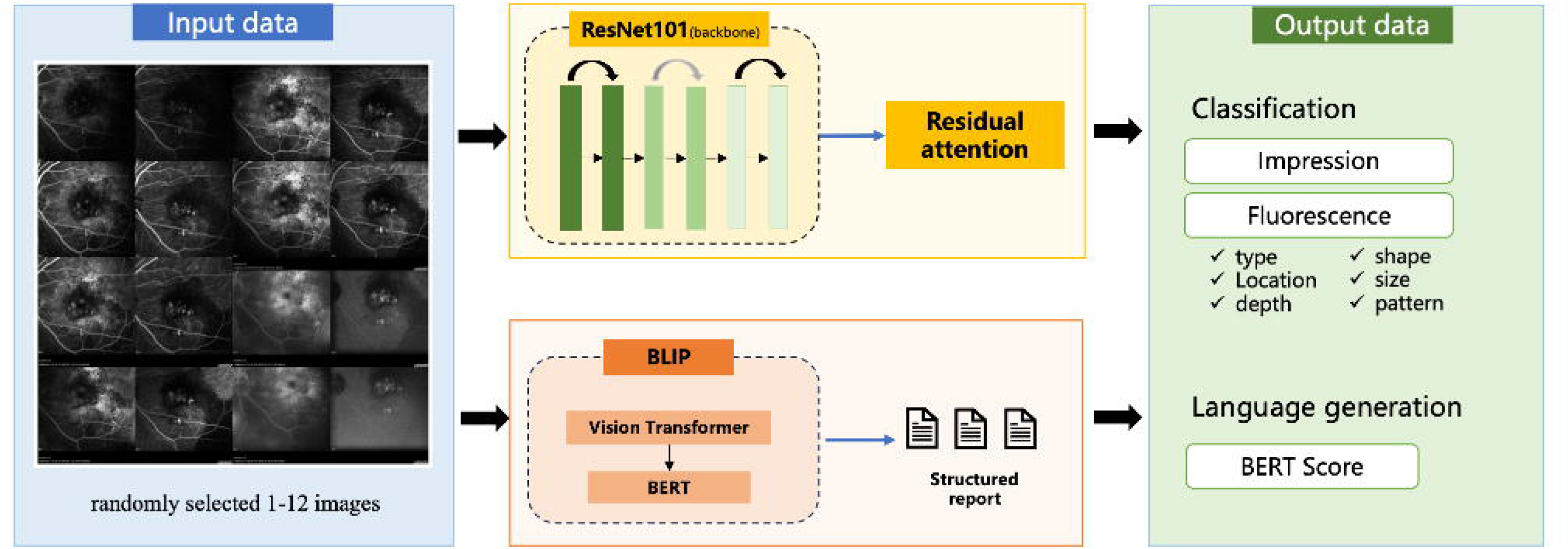
Demonstration of classification and generation methods.

Classification-based: We used the multi-label classification method incorporating class-specific residual attention [14], with ResNet101[15] as the backbone. This method proves particularly effective for the intricate task of multi-label image recognition. Leveraging its simplicity and efficiency, class-specific residual attention captures distinct spatial regions occupied by objects from angiographic lesion different categories. During training, we randomly selected 1-12 angiographic images from each case for model input, leveraging pretrained weights from ImageNet. The angiography images were resized to 384×384, and we employed the Adam optimizer[16] during the fine-tuning process. The initial learning rate was set at 0.01, accompanied by a weight decay of 0.0001 and a batch size of 4. The model with the highest mean average precision on the validation set was selected for testing.

Language-generation based: We utilized the Bootstrapping Language-Image Pre-training (BLIP) framework[17] for our approach. This framework allows for vision-language pre-training and subsequent fine-tuning of the model on report generation tasks. The framework incorporates a vision transformer[18] as the image encoder(BERT)[19] as the language encoder and decoder, and a captioner to filter out noisy training data. After extensive pre-training on a large corpus of language and image data, BLIP has demonstrated impressive generalization capabilities in image captioning, effectively bridging the language and vision modalities.[17] For our angiographic report generation task, we fine-tuned the pre-trained model using FFA and/or ICGA images along with structured reports. During training, consistent with the classification method, we randomly selected 1-12 images from each case for model input. The images were resized to 384×384, and the AdamW optimizer[16] was used during the fine-tuning process. The initial learning rate was set at 0.00002, with a weight decay of 0.05, a batch size of 4, and a cosine learning rate schedule. Model selection for testing was based on achieving the highest Bilingual Evaluation Understudy (BLEU1)[20] score on the validation set. BLEU assesses position-independent sequential matching by comparing the n-gram candidates with the n-gram references, which assesses the precision of n-grams (up to 4-grams) in the generated text relative to the reference text, providing a refined evaluation in the context of angiographic report generation.

Model training for both classification and language generation methods was conducted for 30 epochs using an NVIDIA Tesla V100 GPU.

### Evaluation

We conducted a comprehensive assessment of our report-generation solutions using both classification and language-generation method metrics.

Classification metric: The evaluation was based on the F1 score, chosen for its resilience against imbalances in data distribution across 14 different classification tasks targeting various features and lesions. Firstly, we determine the overall impression of each eye. For hyperfluorescence assessment, we identify the hyperfluorescence type (HyperF_Type) and measure the hyperfluorescence area (HyperF_Area) in disc areas (DA). The lesion can be classified as -4 if it is 4 or smaller than 4 disc areas, or as -5 if larger than 4 disc areas. We also identify hyperfluorescence in the fovea (HyperF_Fovea), outside the fovea (HyperF_ExtraFovea), and at the Y location (HyperF_Y). Similarly, for the hyperfluorescence tasks, we determine the type (HypoF_Type), area (HypoF_Area) in disc areas, in the fovea (HypoF_Fovea), outside the fovea (HypoF_ExtraFovea), and at the Y location (HypoF_Y). Furthermore, we detect choroidal neovascularization (CNV) and vascular abnormalities related to diabetic retinopathy (DR). Finally, we analyze the pattern of the observed abnormalities.

For each subtask, the evaluation was based on the average F1 score according to the following formula:

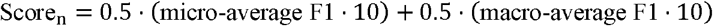

1. Micro-average F1 Score
  - Precision and Recall were calculated for each class.
  - The micro-average F1 score, representing an aggregate measure across all classes, was calculated using the formula:

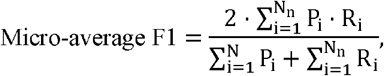

where N_*n*_ is the total number of classes in subtask *n*.
2. Macro-average F1 Score
  - Precision and Recall were calculated for each class.
  - The F1 score for each class was computed using the formula:

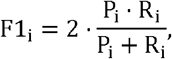

The macro-average F1 score, representing an average across all classes, was calculated as: :

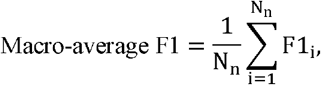

where N_*n*_ is the total number of classes in subtask *n*.

In each subtask, where samples may belong to different categories or have multiple labels, the final subtask score combined micro and macro F1 scores, scaled by a factor of 10 for clarity. This approach provides a holistic evaluation of model performance, addressing sensitivity to class distribution and offering insights into specific class performance, which is well-suited to our clinical scenario.

Language generation metric: To ensure that the classification results align with ground truth at the semantic level, we generated textual descriptions of the reports in the format of “Class name 1: Answer 1, Class name 2: Answer 2…”. We used BERTscore[21], a widely used measurement standard in natural language processing. BERTscore evaluates the semantic similarity between generated and reference sentences using context embedding, providing a refined assessment compared to traditional indicators such as BLEU. Consistent with the classification task, the BERTscore was also magnified by a factor of 10.

## Results

### Dataset

After excluding 226 eyes with media opacity that prevented fundus visualization for diagnosis, the dataset consisted of 55361 images from 1691 patients (3179 eyes). The majority (81.8%) of these eyes were examined in both FA and ICGA modes. Venous phase was the most common (52.4%), followed by late phase (44.4%) and arterial venous phase (3.2%). Each eye has a median of 28 angiographic images, with an interquartile range of 32. The minimum number of images for an eye is 1, while the maximum is 286. The most prevalent image shape was (868, 1536) (90.6%), followed by (1636, 1536) (6.2%), (868, 768) (2.6%), (612, 1024) (0.5%), and (1124, 1024) (0.0%). **Figure 2** illustrates each image type.

The 3179 eyes were evenly distributed between left (50%) and right (50%) eyes. A total of 24 conditions were identified. Macular neovascularization was the most common, found in 32.5% of the eyes. Unremarkable changes were noted in 21.8% of the eyes. Dry age-related macular degeneration was present in 10.2% of the eyes, while central serous chorioretinopathy was found in 9.0%. Uveitis and chorioretinal scar were each found in 4.6% of the eyes. Diabetic retinopathy was observed in 3.2% of the eyes, retinal pigment epithelial detachment in 2.8%, polypoidal choroidal vasculopathy in 2.4%, and pachy choroid pigment epitheliopathy in 2.1%. Less common conditions included chorioretinal atrophy (1.0%), myopia (0.8%), proliferative diabetic retinopathy (0.8%), cystoid macular edema (0.7%), and choroidal mass (0.7%). Other conditions, including epiretinal membrane, retinal vein occlusion, and others, were noted in 0.5% of the eyes. The least common conditions were retinal arterial macroaneurysm (0.3%), branch retinal vein occlusion (0.2%), central retinal vein occlusion (0.2%), retinal dystrophy (0.2%), diabetic macular edema (0.2%), and central retinal artery occlusion (0.1%).

Hyperfluorescence manifested in various forms, with leakage being the most common type (39.5%), followed by staining (19.9%), no hyperfluorescence (24.4%), pooling (9.8%), and window defect (6.4%). The hyperfluorescence area was most commonly 4 disc areas (61.2%), with 14.4% having 5 disc areas and 24.4% showing no hyperfluorescence. Hyperfluorescence was absent in the fovea in 51.2% of cases, while it was present in 48.8% of cases. Hypofluorescence was observed as a blockage in 24.3% of cases, capillary non-perfusion in 3.8% of cases, and was absent in 71.9% of cases. The hypofluorescence area was 4 disc areas in 20.6% of cases, 5 disc areas in 7.5% of cases, and was absent in 71.8% of cases. Hypofluorescence was absent in the fovea in 80% of cases, while it was present in 20% of cases. Choroidal neovascularization (CNV) was present in 34.4% of cases. Other multilabel manifestations of fluorescence are presented in **Supplementary Table 1**.

### Baseline Performance

The performance of our baseline models is presented in **Table 2**. The classification method achieved an average score of 7.966, which is a composite of the overall F1 score (5.977) and a BERT score (9.955). For the F1 score of lesion/impression/pattern recognition tasks, the model achieved an impression score of 3.795, a CNV score of 8.164, a vascular abnormality (DR) score of 5.684, and a pattern score of 5.673. For hyperfluorescence metrics, the model achieved a type score of 4.731, an area (DA) score of 7.627, a fovea score of 7.683, an extra fovea score of 3.261, and an Y direction score of 6.639. For hypofluorescence metrics, the classification method achieved a type score of 6.559, an area (DA) score of 6.294, a fovea score of 7.236, an extra fovea score of 4.468, and an Y direction score of 5.857.

**Table 2.** Model performance on the test set.

|  | Overage score | F1score | BERT score | Impression score | Hyperfluorescence type score | Hyperfluorescence area (DA) score | Hyperfluorescence fovea score | Hyperfluorescence extra fovea score | Hyperfluorescence (y) score |
| --- | --- | --- | --- | --- | --- | --- | --- | --- | --- |
| Classification method | 7.966 | 5.977 | 9.955 | 3.795 | 4.731 | 7.627 | 7.683 | 3.261 | 6.639 |
| Language generation method | 7.947 | 5.939 | 9.956 | 4.505 | 5.317 | 6.963 | 7.656 | 3.498 | 6.458 |
|  |  | Hypofluorescence type score | Hypofluorescence area (DA) score | Hypofluorescence fovea score | Hypofluorescence extra fovea score | Hypofluorescence (y) score | CNV score | Vascular abnormality (DR) score | Pattern score |
| Classification method |  | 6.559 | 6.294 | 7.236 | 4.468 | 5.857 | 8.164 | 5.684 | 5.673 |
| Language generation method |  | 6.529 | 5.79 | 7.116 | 4.361 | 5.614 | 7.781 | 5.676 | 5.878 |

The language generation method demonstrated comparable performance with an average score of 7.947, which is a composite of the overall F1 score of 5.939 and a BERT score of 9.956. For the F1 score of lesion/impression/pattern recognition tasks, the language generation method achieved an impression score of 4.505, a CNV score of 7.781, a vascular abnormality (DR) score of 5.676, and a pattern score of 5.878. For hyperfluorescence metrics, the method achieved a type score of 5.317, an area (DA) score of 6.963, a fovea score of 7.656, an extra fovea score of 3.498, and an y-direction score of 6.458. For hypofluorescence metrics, it achieved a type score of 6.529, an area (DA) score of 5.79, a fovea score of 7.116, an extra fovea score of 4.361, and a y score of 5.614.

Both methods exhibit very similar overall performance, with the classification method having a slightly higher average score. The classification method outperforms in CNV recognition, while the language generation method performs better in impression recognition. For hyperfluorescence tasks, the classification method performs better in hyperfluorescence type and area recognition, while the language generation method excels in fovea recognition. Both methods show comparable performance in hypofluorescence metrics, with similar scores across the different tasks. In summary, the classification method and the language generation method exhibit comparable overall performance, with each method showing strengths in specific recognition tasks.

## Discussion

The automatic interpretation of angiographic reports is crucial for aiding medical decision-making. In this study, we have released the largest multimodal angiographic dataset to date and provided two baseline solutions for reference.

The rapid advancement of digital health and artificial intelligence (AI) applications offers an opportunity to revolutionize eye health. This can be achieved by facilitating access to eye care and supporting clinical decision-making through an objective, data-driven approach. While several studies have explored the use of FFA images for automatic report generation[8, 10, 22], it has been reported only 52% of the 204 reported ophthalmic databases are available online.[23] Furthermore, there are no publicly available large angiographic datasets for research purposes. Our study addresses this gap by proposing a large, well-curated angiographic dataset with detailed labels.

We have considered the granularity and logic of the label from a clinical perspective, ranging from the overall impression to delicate fluorescein changes, as well as the location and depth of these changes. This level of detail has not been achieved in previous studies. Competitors can utilize these medically meaningful labels through multi-label classification, report generation methods, or hierarchical image classification to generate a structured report.

However, the dataset presents a significant challenge as it was collected from a real clinic without biased selection. Therefore, the conditions in the dataset are diverse and skewed, with a majority of common conditions and some rare cases. Therefore, we provided two baseline methods: one purely for classification and another for language generation. We used a single model for all tasks, assuming that the tasks would complement each other. Competitors are free to divide the 14 tasks and optimize each one individually. Additionally, class weight could be added to improve the performance of the metric for specific tasks.

## Conclusion

We have curated the largest multimodal angiographic data with detailed labels for report generation. Both classification and report generation methods can achieve acceptable performance. We hope our approach will significantly accelerate the application of AI in ophthalmology.

## Supporting information

Supplemental Table 1. Multilabel manifestations of fluorescence

## Data Availability

All data produced in the present study are available upon reasonable request to the authors.

## Abbreviations

FFA: Fundus Fluorescein Angiography
CF: Color Fundus Photography
OCR: Optical Character Recognition

## Authors’ contributions

D.S. and P.R. conceived the study. P.C. collected and labeled data. D.S. and P.C. cleaned the data. W.Z. built the deep learning model. W.Z. and P.C. did the literature search, analyzed the data, and wrote the manuscript. W.Z., D.S., P.C. P.R., and M.H. contributed to key data interpretation. All authors critically revised the manuscript.

